# Cerebrospinal fluid and plasma biomarkers in individuals at risk for genetic prion disease

**DOI:** 10.1101/2019.12.13.19014217

**Authors:** Sonia M Vallabh, Eric Vallabh Minikel, Victoria J Williams, Becky C Carlyle, Alison J McManus, Chase D Wennick, Anna Bolling, Bianca A Trombetta, David Urick, Chloe K Nobuhara, Jessica Gerber, Holly Duddy, Ingolf Lachmann, Christiane Stehmann, Steven J Collins, Kaj Blennow, Henrik Zetterberg, Steven E Arnold

## Abstract

**BACKGROUND:** Fluid biomarkers are important in the development of therapeutics to delay or prevent prion disease, but have not been systematically evaluated in pre-symptomatic individuals at risk for genetic prion disease.

**METHODS:** We recruited pre-symptomatic individuals with pathogenic mutations in the prion protein gene (*PRNP*; N=27) and matched controls (N=16), to donate cerebrospinal fluid (CSF) and blood at multiple timepoints to a cohort study at Massachusetts General Hospital. We quantified total prion protein (PrP) and real-time quaking-induced conversion (RT-QuIC) prion seeding activity in CSF, and the neuronal damage markers total tau (T-tau) and neurofilament light chain (NfL) in both CSF and plasma. We compared these markers cross-sectionally between mutation carriers and controls, evaluated short-term test-retest reliability over 2-4 months, and conducted a pilot longitudinal study over 10-20 months for a subset of participants.

**FINDINGS:** CSF PrP levels were stable on test-retest with a mean coefficient of variation of 7% both over 2-4 months in N=29 participants and over 10-20 months in N=10 participants. RT-QuIC was negative in 22/23 mutation carriers. The sole individual with positive RT-QuIC seeding activity at two study visits had steady CSF PrP levels and slightly increased tau and NfL concentrations compared with others, though still within the normal range, and remained asymptomatic one year later. Overall, tau and NfL showed no significant differences between mutation carriers and controls in either CSF or plasma.

**INTERPRETATION:** CSF PrP will be interpretable as a pharmacodynamic readout of the effects of a PrP-lowering therapeutic in pre-symptomatic individuals, and may serve as a surrogate biomarker in a “primary prevention” trial paradigm. In contrast, current markers of prion seeding activity and of neuronal damage do not reliably cross-sectionally distinguish mutation carriers from controls, arguing against the feasibility of a “secondary prevention” paradigm in which trials specifically recruit pre-symptomatic participants with prodromal evidence of pathology.

## BACKGROUND

The well-defined pathobiology of prion disease, with prion protein (PrP) as the sole causal agent^1^, has spurred the preclinical development of rational therapeutics to lower PrP expression in the brain^2^. However, the rapid progression of prion disease, which is typically fatal in under a year^3^, presents a challenge for drug development, as symptomatic patients may be profoundly debilitated by the time of diagnosis and enrollment^4,5^. Individuals with mutations in the prion protein (*PRNP)* gene, many of which are highly penetrant^6^, may be aware of their risk decades in advance of symptom onset^7^, creating an opportunity for early intervention for primary prevention. However, as the highly variable and unpredictable age of onset precludes randomizing individuals to a treatment arm for years or decades until manifest disease^8^, surrogate biomarker trial endpoints will be essential for drug development.

In “secondary prevention” paradigms, trials may recruit pre-symptomatic individuals with prodromal biomarker evidence of a disease process underway and aim to stabilize or reverse progression of those pathological biomarkers^9^. This approach may be useful in slowly progressive dementias including Alzheimer’s disease and Huntington’s disease thanks to decades-long phases of prodromal pathology during which imaging and fluid biomarkers are progressive^10,11^ To date in prion disease, however, longitudinal imaging and neurophysiological studies have uncovered at most subtle changes in a small number of individuals only around one year before disease onset^12–14^, and systematic characterization of fluid biomarkers in pre-symptomatic individuals at risk has not been described.

One proposed primary prevention paradigm for prion disease involves lowering native PrP levels in the brain, a goal in principle achievable by targeting the prion protein gene, its RNA, or the PrP protein product prior to misfolding^15^. In this scenario, reduction of the causative fuel for pathogenic prion formation prevents or delays its ignition into a fulminant disease. Pre-symptomatic individuals at known high genetic risk but with no requirement of prodromal pathology could be treated and a therapeutic could be evaluated based solely on a pharmacodynamic biomarker. In this case, CSF PrP is a promising biomarker candidate given the absolute necessity of PrP to the disease process^15^. To enable development of such primary preventions, it is essential to be able to measure PrP precisely and to understand the natural intra-individual variability of PrP levels in pre-symptomatic individuals. We previously described the technical precision and pre-analytical factors that might affect measurements of PrP levels in CSF, as well as their biotemporal stability within non-mutation carrying individuals with mild cognitive impairment over the short term (approximately 8 weeks)^16^.

Here we set out to recruit and characterize a cohort of pre-symptomatic individuals at risk for genetic prion disease, and controls, some of whom we have followed for over a year. Our goal was to evaluate the within-subject stability of PrP in CSF fluid biomarkers and to characterize other emerging fluid biomarkers of neurodegeneration in pre-symptomatic at-risk individuals for their potential utility in either primary or secondary prevention paradigms.

## RESULTS

Individuals recruited to the study included (see Methods): 1) confirmed mutation carriers of *PRNP* variants, 2) untested individuals at risk of carrying a *PRNP* variant, usually established through confirmed mutation status of a first-degree relative and a family history of autopsy-proven prion disease; and 3) non-carrier controls. All participants were genotyped to establish *PRNP* mutation carrier status. Mutation carriers and controls were demographically well-matched (Table 1). All participants scored 20/20 points at each visit on the prion disease-specific MRC functional rating scale^17^, and carriers performed comparably to controls on the Montreal Cognitive Assessment^18^. In total, a battery of 20 cognitive, neuropsychological, psychiatric and motor tests and inventories established that all participants performed within established normative ranges, with no significant differences between mutation carriers and controls, supporting the carriers’ pre-symptomatic status (Table S1). The research lumbar puncture (LP) was well-tolerated, and the N=24 participants for whom this was the first LP generally reported lower anxiety about undergoing future LPs than they had felt about the first LP (Figure S1).

**Table 1:** Demographic overview of study participants. Participant number, age, sex, PRNP genotype, total number of study visits at time of analysis, and scores on two basic assessments of daily and cognitive functioning. Corrected P values account for all 20 assessments performed. The “other” category includes four distinct mutations, two of which are of low penetrance and two of which are highly penetrant^6,8^; to protect participant confidentiality, the exact mutations are not disclosed.

|  |  | *PRNP*<br>mutation<br>carriers | Non-carrier controls |  |  |
| --- | --- | --- | --- | --- | --- |
| N |  | 27 | 16 |  |  |
| Age at first visit |  | 44.2±15.2 | 44.5±12.7 |  |  |
| Sex | Male | 10 | 5 |  |  |
|  | Female | 17 | 11 |  |  |
| *PRNP*<br>genotype | Wild type | 0 | 16 |  |  |
|  | E200K | 12 | 0 |  |  |
|  | D178N | 7 | 0 |  |  |
|  | P102L | 4 | 0 |  |  |
|  | Other | 4 | 0 |  |  |
| Number of<br>completed<br>study visits | 1 visit | 7 | 4 |  |  |
|  | 2 visits | 9 | 9 |  |  |
|  | 3 visits | 11 | 3 |  |  |
|  |  | *PRNP*<br>mutation<br>carriers | Non-carrier<br>controls | P value | P value<br>(Bonferroni<br>corrected) |
| Assessments | MRC prion<br>disease rating<br>scale | 20.0±0.0 | 20.0±0.0 | 1 | 1 |
|  | Montreal<br>cognitive<br>assessment | 27.7±1.6 | 28.5±1.7 | 0.20 | 1 |

### Within subject test-retest stability of CSF PrP

To confirm the short-term within-subject stability of CSF PrP levels in genetic prion disease mutation carriers and controls, we quantified PrP levels in CSF samples given by study participants at 2-4 month intervals using an Enzyme Linked Immunosorbent Assay (ELISA) that we have previously shown appropriate for this purpose^16^. Following collection, CSF used for this analysis was handled uniformly according to an established protocol that includes addition of 0.03% CHAPS detergent to maintain PrP solubility^16^. CSF PrP levels were stable over this interval and similar between carriers (mean CV = 6.8%) and controls (mean CV = 7.5%) (Figure 1A). CSF PrP concentration showed a significant difference between genotypes (p=0.016, one-way ANOVA) driven by lower PrP in D178N mutation carriers (Figure 1A; see discussion). While all reasonable efforts were made to standardize CSF collection, in some cases clinical variations were noted, including use of drip collection rather than aspiration and lower sample yields. On average, the six individuals whose CSF was handled differently between the two visits showed greater, though still reasonable, variation in CSF PrP levels (mean CV = 12.6%) compared to all other participants (mean CV = 5.8%).

**Figure 1:**
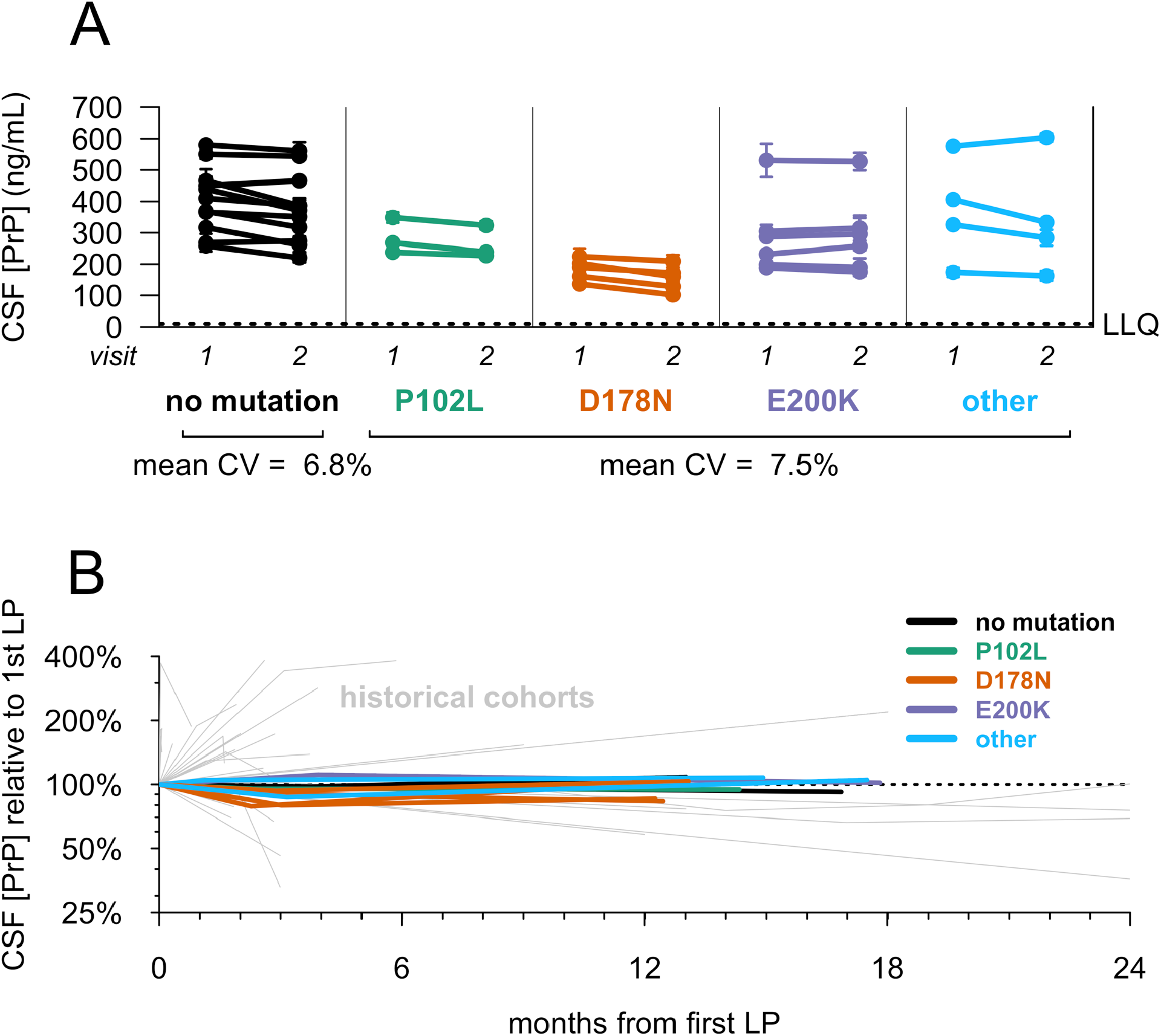
Test-retest stability of CSF PrP. Uniformly processed CSF samples were collected from lumbar punctures performed by one of two investigators (SEA, AJM). CSF PrP levels were quantified by ELISA. Dots represent means, and line segments 95% confidence intervals, of measurements within dynamic range with 2 technical replicates each. A. 29 individuals gave two CSF samples at an interval of 2-4 months. B) Ten participants with the noted genotypes gave three CSF samples at the following intervals: initial visit, 2-4 month follow-up visit, 10-20 month follow-up visit. For each subject, PrP levels for all visits have been normalized to levels at the first visit, such that the first LP is defined as 100%. Grey lines show PrP test-retest stability for CSF samples from previously reported retrospective cohorts without uniform sample handling to minimize pre-analytical variability, reproduced from Vallabh et al^16^.

At the time of analysis, ten individuals had completed a longitudinal study visit 10-20 months after their initial study visit, enabling a longer-term analysis of CSF PrP test-retest stability. Across eight mutation carriers and two controls, CSF PrP levels were again steady with a mean CV of 7.2% across all three visits (Figure 1B). This variability is far lower than we have previously observed in test-retest CSF samples from retrospective cohorts lacking uniform sample handling^16^ (Figure 1B), consistent with pre-analytical factors being a major source of variability in those cohorts. Across all participants, CSF PrP was modestly correlated with CSF total protein (r=0.35, p=0.0052, two-sided Spearman’s correlation), replicating previous reports^16,19^.

### Disease process biomarkers in CSF

To assess levels of emerging markers of neuronal damage or neurodegeneration in carrier and control CSF, we measured total tau (T-tau) and neurofilament light (NfL) levels in participant CSF by ELISA. Uniformly handled CSF aliquots with no additive were used for these assays and all CSF assays that follow. Both CSF T-tau and CSF NfL were highly elevated in positive control CSF from symptomatic prion disease patients (p=2.6 × 10^−10^ for T-tau; p=1.6 × 10^−11^ for NfL, 2-sided Kolmogorov-Smirnov test; Figures 2A-B). In contrast, levels of these markers in mutation carrier and non-carrier control CSF were similar. CSF T-tau appeared nominally higher in non-carrier controls (mean 251.8±84.5 pg/mL) than in mutation carriers (224.5±112.3 pg/mL) (uncorrected p=0.015, 2-sided Kolmogorov-Smirnov test), while CSF NfL was indistinguishable between the two groups (p=0.61, 2-sided Kolmogorov-Smirnov test). CSF T-tau and NfL were independently measured by ELISA at a second site, and values showed good correlation with those values initially obtained (T-tau, r=0.80, p=3.5 × 10^−9^; NfL, r=0.89. p=3.0 ×10^−14^), again with no difference between carriers and controls (Figure S2).

**Figure 2:**
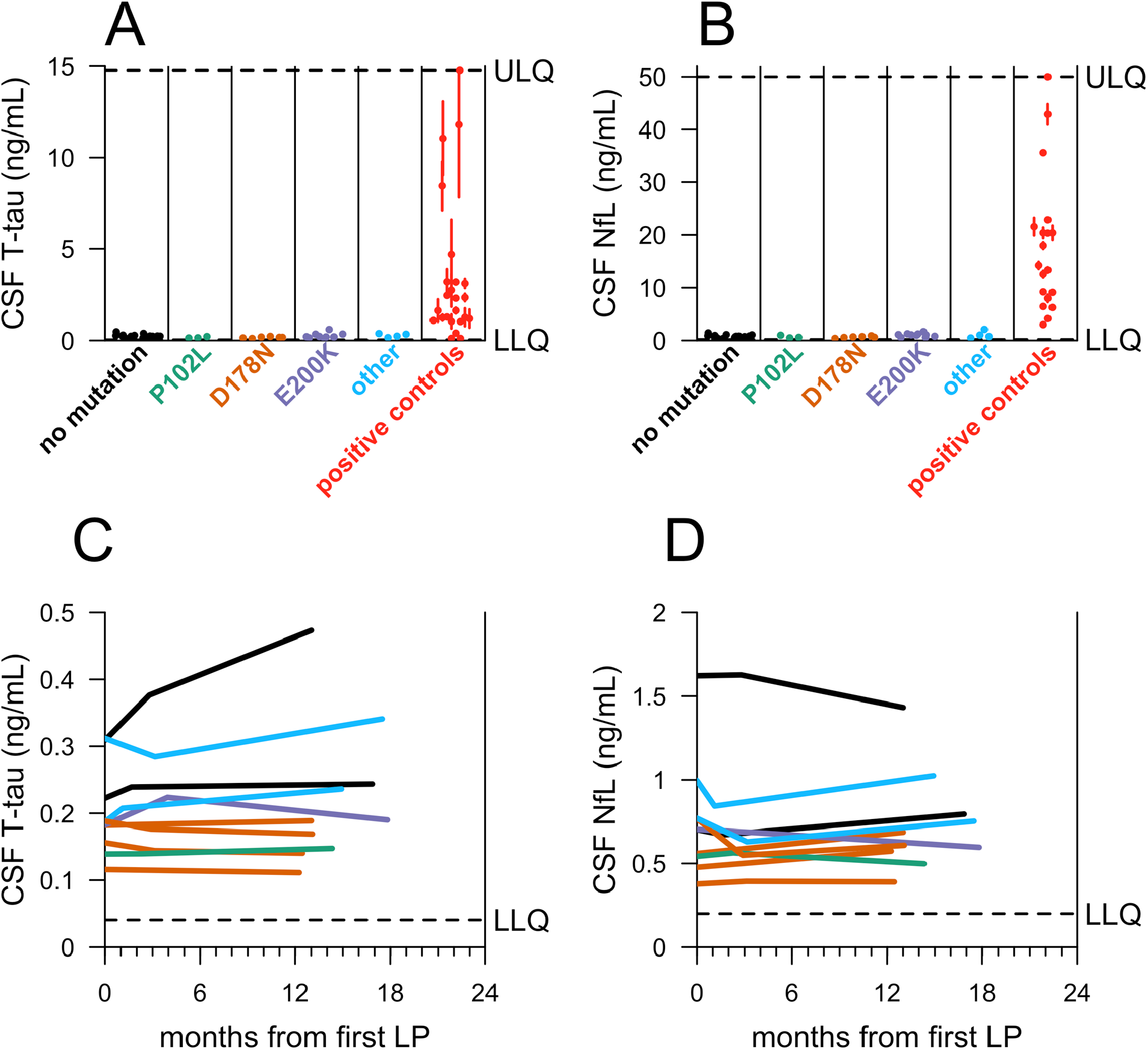
Candidate markers of neuronal damage in carrier and control CSF (Broad Institute). CSF **A)** T-tau and **B)** NfL levels were measured by ELISA for N=39 participants who have made at least one study visit, for whom genotypes were available at time of analysis, and where appropriate CSF aliquots were available. For each participant included, samples were taken from the most recent visit at time of analysis. Symptomatic prion disease CSF samples (red, N=24 for T-tau, N=19 for NfL) were included from both sporadic and E200K genetic prion disease. The operator was masked to mutation status. Dots represent means, and line segments 95% confidence intervals, of measurements within dynamic range with 2 technical replicates each. **C and D)** For N=10 participants who had completed a longitudinal visit ≥10 months after their first visit, both T-tau and NfL were measured by ELISA across all visits to assess longitudinal dynamics. As in Figure 1B, CSF from the following three timepoints is represented for each participant: initial visit, 2-4 month follow-up visit*, 10-20 month follow-up visit. (*The 2-4 month follow-up visit was missing for N=1 participant in **C** and **D**).

To assess potential changes over time in markers of neuronal damage, we quantified total T-tau and NfL in the longer-term serial CSF samples available for the N=10 longitudinal participants spanning 10-20 months by ELISA. Across all visits (10-20 months), levels of both proteins remained low with no significant change over time within individuals (p=0.51 for T-tau, p=0.91 for NfL, linear regression; Figures 2C-D). The mean CV over all visits was 7.8% for CSF T-tau and 9.9% for CSF NfL.

Recent reports have demonstrated that in addition to being detectably elevated in the CSF of symptomatic prion disease patients, T-tau and NfL are also elevated in blood^20–23^. Levels of both analytes were therefore measured in participant plasma using the Simoa platform^21,24^. Plasma T-tau measurements showed a relatively high level of within-subject fluctuation over a short time frame (mean CV = 38%), limiting interpretability (Figure S3A). Plasma NfL, on the other hand, showed better within-subject reliability over the same interim of 2-4 months (mean CV = 18%; Figure S3B). Group-wise, plasma NfL levels were not significantly different between controls and carriers (p=0.46, two-sided Kolmogorov-Smirnov test; Figure 3A), and all individuals were within normal ranges, well below the typical values reported in symptomatic genetic prion disease patients^20,21^. We observed no temporal trend in plasma NfL among participants who made three visits over 10-20 months (p=0.91, linear regression, Figure 3B).

**Figure 3:**
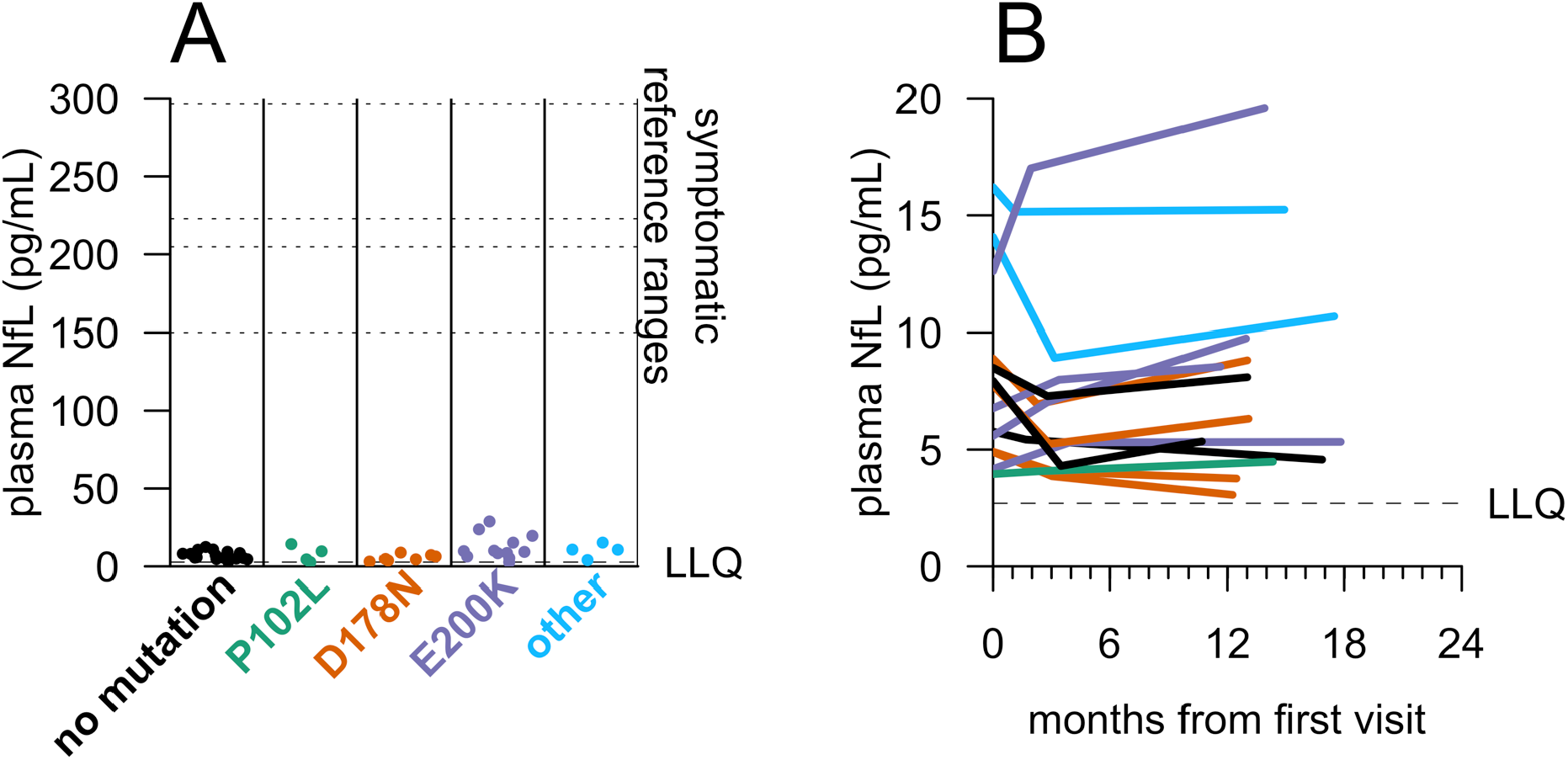
NfL levels in carrier and control plasma. Plasma NfL levels were measured by Quanterix Simoa assay. A) For N=43 participants who have made at least one study visit, samples were taken from the most recent visit at time of analysis. Dots represent singlicate measurements. Dashed lines for “symptomatic reference ranges” represent mean values reported for symptomatic genetic prion disease patients^20,21^ or median values reported for symptomatic sporadic prion disease patients^22,23^. **B)** For N=14 participants who had completed three visits, plasma NfL levels were measured by Quanterix Simoa for all three visits to assess longitudinal dynamics. As in Figure 1B, CSF from the following three timepoints is represented for each participant: initial visit, 2-4 month follow-up visit, 10-20 month follow-up visit.

We measured prion seeding activity in participant CSF samples, using the real-time quaking induced conversion (RT-QuIC) assay now in widespread clinical diagnostic use^25^. Using established pre-specified criteria^25^, positive reactions were defined as those where at least 2/4 replicates reached a predetermined fluorescence threshold within 24 hours of initiation of the assay. Under these conditions, positive controls including CSF from both sporadic and genetic post-mortem confirmed prion disease cases were identified as positive with 88% sensitivity. This is comparable to reported results^25–27^ (Figure 4A). Non-carrier control samples were negative (Figure 4B), as were 22/23 carrier samples (Figures 4C-4F). One E200K mutation carrier had a positive result (Figure 4E) and is discussed in greater detail below.

**Figure 4:**
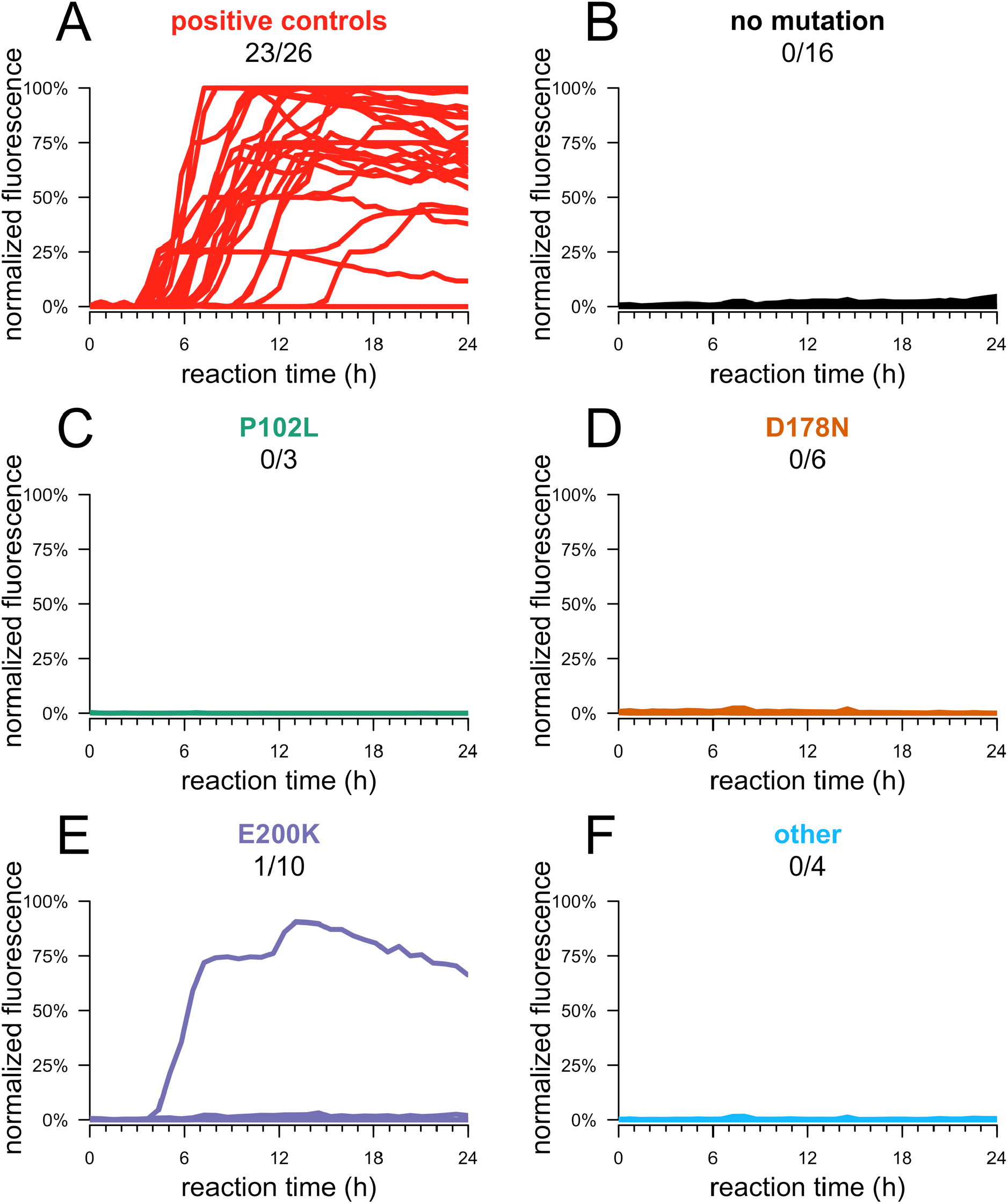
RT-QuIC seeding activity in carrier and control CSF, using SHaPrP substrate. RT-QuIC was performed on CSF from 39 participants who made at least one study visit. For each participant, samples were taken from the most recent visit at time of analysis. RT-QuIC was performed following an established protocol for second-generation CSF RT-QuIC using SHaPrP substrate. Reactions were seeded with 20μL CSF from N=26 symptomatic prion disease cases and N=39 MGH study participants, with each reaction run in quadruplicate.

#### Kinetic curves – normalized thioflavin T (ThT) fluorescence (y axis) vs. time in hours (x axis) – are shown for each sample, averaged across four replicates

Because RT-QuIC has shown more limited sensitivity for symptomatic genetic prion disease CSF under standard conditions^26,27^ than sporadic prion disease, we also tested recombinant bank vole prion protein (BvPrP), which was reported as a seemingly universal substrate for RT-QuIC detection of diverse prion strains in brain homogenate^28^, though its utility for CSF has not been explored. Per published criteria^28^, positive samples were those for which at least 2/4 replicates reached a predetermined fluorescence threshold within 50 hours of initiation of the assay. With these conditions and criteria, this assay exhibited nominal, albeit weak positives in 10/19 (53%) of positive control CSF samples from symptomatic prion disease cases, and spontaneous reactions in 2/16 mutation-negative controls (Figure S4), limiting interpretability of this version of the assay.

### RT-QuIC positive pre-symptomatic carrier

One participant, a carrier of the E200K mutation of age *≥*60, showed RT-QuIC seeding activity upon analysis of CSF from their second visit. We subsequently performed additional analysis in order to fully assess RT-QuIC seeding activity, CSF T-tau and NfL levels, and plasma NfL levels, from both visits 1 and 2 for this individual. 4/4 RT-QuIC replicates were positive at both visits. CSF NfL remained in the normal range, comparing both to within-study normal controls and published reference ranges^20,21^. CSF T-tau and plasma NfL were modestly higher, for both visits, than controls and other mutation carriers in our study, consistent with age (Table 2; see Discussion). This participant’s score on the MRC prion disease rating scale remained stable. While their score on the Montreal Cognitive Assessment (MoCA) declined nominally between visits 1 and 2, from 27 to 25 out of a possible 30, fluctuation of one to three points was common between first and second visits in our participants, with differences of up to six points noted; furthermore, ten other participants scored 25 or less on the MoCA at least once over their first two visits. A full battery of 20 tests spanning cognitive, psychiatric, motor and daily living assessments revealed no striking or consistent changes (Table S2). This individual remained asymptomatic >1 year after their second visit. CSF PrP levels were in the middle of the observed range, and were stable between visits 1 and 2.

**Table 2:** Comparison of visits for one RT-QuIC-positive study participant. RT-QuIC replicates were designated as positive based on criteria described above, in methods, and elsewhere^25^. CSF T-tau, NfL, and PrP were measured by ELISA as described in Figures 1 and 2. Plasma NfL was measured by Simoa as described in Figure 3.

|  |  | **Visit 1** | **Visit 2** |
| --- | --- | --- | --- |
| **Fluid biomarkers** | CSF PrP (ng/mL) | 287 | 296 |
|  | CSF T-tau (ng/mL) | 0.57 | 0.60 |
|  | CSF NfL (ng/mL) | 1.27 | 1.48 |
|  | Plasma NfL (pg/mL) | 23.5 | 28.8 |
|  | RT-QulC (positive replicates) | 4/4 | 4/4 |
| **Assessments** | MRC prion disease rating scale | 20 | 20 |
|  | Montreal Cognitive Assessment | 27 | 25 |

## DISCUSSION

Here we describe interim results from an ongoing longitudinal clinical study characterizing genetic prion disease mutation carriers and mutation-negative controls. We evaluate candidate fluid biomarkers for primary and secondary prevention trial designs in pre-symptomatic genetic prion disease.

PrP-lowering therapeutics are now in preclinical development, so CSF PrP will be important as a pharmacodynamic biomarker at a minimum, and regulators have expressed openness to its use as a surrogate endpoint in pre-symptomatic individuals^15^. Productive use of this marker in trials, however, will require dependable performance, including a stable baseline, which might not exist if CSF PrP exhibits high biotemporal variability or if pre-symptomatic individuals exhibit decline in CSF PrP, similar to the lowered CSF PrP levels seen in symptomatic prion disease patients^16,29–31^. Our findings address both of these concerns.

Most historical cohorts in which CSF PrP has been evaluated exhibit large variability in CSF PrP levels^16,29–31^, but we previously reported tight 8-11 week test-retest reliability (mean CV=13%) among uniformly handled samples, suggesting that PrP can be reliably measured if pre-analytical factors are well controlled. Here, we validate this hypothesis prospectively, observing a mean CV of only 7% over 10-20 months among samples handled with exquisite uniformity and early addition of detergent to minimize adsorption to plasticware. This level of biotemporal stability is comparable to that reported for core CSF biomarkers including amyloid beta (Aβ) 1-38, Aβ 1-40, T-tau, and NfL over a similar term^32^. In addition, whereas our previous report was based on patients suffering from non-prion mild cognitive impairment^16^, in whom progressive reduction of CSF PrP due to disease would not be expected, here we confirm that CSF PrP concentration is stable even among individuals at high risk of developing prion disease in their lifetimes.

PrP levels in the CSF of symptomatic D178N individuals have been reported to be lower than in individuals with other *PRNP* mutations or no mutation^31^, a finding replicated here among pre-symptomatic carriers. This difference has been interpreted by some to indicate a prodromal disease process underway^31^. However, other reports suggest that preferential degradation of D178N mutant PrP may occur constitutively, independent of initiation of the disease process^33–36^, leading to lower baseline PrP levels in carriers of this mutation. Our data support the latter interpretation. Relatively lower CSF PrP levels in D178N carriers appear to be stable over short (Figure 1A) and longer terms (Figure 1B), suggesting that this level is constant rather than a dynamic function of approaching symptom onset, which to date has not been observed for any study participant. Notably, CSF PrP levels were stable even in one E200K mutation carrier with RT-QuIC seeding activity (Table 2). Thus, the decline in CSF PrP levels seen in symptomatic disease likely emerges later in the disease process, and should not confound CSF PrP stability in carriers with no observable pathology.

Broadly, these findings suggest that CSF PrP levels are stable enough in any one individual, regardless of *PRNP* mutation, to informatively report on a PrP-lowering therapeutic such as a PrP-lowering antisense oligonucleotide (ASO) in the central nervous system, over time frames likely to be of relevance to dose-finding and biomarker-based trials. In a Phase I/II trial of the huntingtin-lowering ASO HTTRx (RG6042), mutant huntingtin protein was reduced by a mean of 40% in CSF in the two highest dose cohorts^37^; our data suggest that a similar reduction in PrP levels, if achievable, would be reliably detected in CSF. The LP tolerability data we were able to collect suggests that intrathecal delivery of a drug will not be a barrier to treatment among pre-symptomatic carriers of *PRNP* mutations. While our study is biased toward highly motivated carriers willing to participate in research, this same bias will likely apply to trial recruitment, supporting the relevance of these data.

Across the assays employed, most carriers could not be distinguished from non-carrier controls. Put differently, the present data do not support analogies between the disease state of the average adult carrier and the clinically silent incubation phase of prion disease observed in animals experimentally inoculated or horizontally infected with prions, during which subclinical pathology is evident^38–40^. Both previous cohort studies and case reports have identified anecdotal prodromal imaging and physiological signatures of prion disease in single individuals no more than roughly a year in advance of symptomatic onset^12–14,41–44^. Our findings suggest a similar picture may be true for fluid biomarkers. Prion seeding activity is detectable in 1/23 pre-symptomatic carriers, appearing to pre-date symptom onset in this case by at least one year. While RT-QuIC findings in this individual were clear, with 4/4 replicates positive in both CSF samples tested, biomarkers of neuronal damage in this individual were not abnormal. Plasma NfL and CSF T-tau appeared marginally elevated relative to other participants in our study, but these values still fall within published ranges for age-matched healthy controls^45–47^ and far below the average levels reported for symptomatic genetic prion disease patients^20,21,48^.

To our knowledge, ours is the first report of RT-QuIC seeding activity in the absence of prion disease symptom onset. These data may suggest that for individuals harboring the E200K mutation, RT-QuIC seeding activity may offer an early sign of pathological change, before behavioral or cognitive scales can detect changes in performance. But this finding should be interpreted with caution. One previous report found that an E200K individual converted from RT-QuIC negative to positive between 2 and 4 months after symptom onset^49^. This difference could reflect differences in RT-QuIC protocol employed, or variability in conversion time relative to symptom onset between individuals. Moreover, 9/10 E200K carriers and 13/13 carriers of other mutations were negative by RT-QuIC. Detectable seeding activity is therefore unlikely to be consistently present for a long period prior to onset.

Our study has several limitations. Here we chose to focus on a small set of fluid biomarkers with the best-established association with the prion disease process. Future analyses could explore other potential sentinel markers of prion disease. Our study is relatively young, and our analyses to date provide only short- to medium-term and cross-sectional findings. Moving forward, participants will be seen at annual intervals where feasible, with an eye to enhancing the longitudinal analysis of CSF PrP and enabling longitudinal tracking of pathological biomarkers. Complementary perspectives on the above may be provided by two other efforts, as yet unpublished, to systematically characterize healthy individuals carrying genetic prion disease predisposing mutations: the UK National Prion Monitoring Cohort^17^ and UCSF Early Diagnosis of Human Prion Disease^50^. Age of onset is highly variable in genetic prion disease, we have no means to predict time to onset for any individual carrier in our cohort, and annual hazard for any given individual is low^8^; prior experience suggests that observing even a handful of conversions in a prospective carrier cohort could take between ten and twenty years. For this reason, in the near term a study of this nature is better positioned to report on the state of the average carrier at a given time than on the dynamics of conversion to the disease state.

A strength of our study is that the highly penetrant prion disease-causing mutations most common in the general population^6^ are also those most represented in our cohort. Our characterization may therefore provide a reasonable cross-sectional snapshot of carriers available for recruitment for research or trials. At present, our findings regarding disease stage biomarkers suggests that a “secondary prevention” strategy may not be feasible in genetic prion disease: any prodromal period may be too subtle, too brief, or present in too few individuals at any given time to enable recruitment of a large enough prodromal cohort to enable trials. It remains possible that a fluid biomarker that reliably presages symptom onset further in advance could emerge from further study, particularly in more slowly progressive genetic prion disease variants, allowing subclinical pathology to be tracked in a small cohort of carriers and leveraged toward a secondary prevention trial design. However, our present findings may reflect where the field is likely to stand as therapeutics presently in development approach clinical trials. In this context, pre-symptomatic trials in genetic prion disease may be better served by a primary prevention model based on genetic risk.

## METHODS

### Study design, participants, and ethical approval

The study was conceived and designed with the pre-specified, publicly announced^51^ primary goal of evaluating the test-retest stability of CSF PrP concentration in individuals at risk for genetic prion disease. This study was approved by the Partners Institutional Review Board in April 2017 (protocol #2017P000214). Participants were recruited through IRB-approved advertisements hosted by Massachusetts General Hospital and Prion Alliance. News of the study was often shared by word of mouth in the prion disease community or by social media accounts run by Prion Alliance and the CJD Foundation. Participants included known mutation carriers, individuals at risk (typically 50/50 risk with an affected first-degree relative), and controls including genetic prion disease family members who had already tested negative for a mutation, spouses, and unrelated but demographically matched local controls. Participation required two study visits to Boston and the absence of any contraindication to lumbar puncture (LP). All participants were cognitively sound and provided written informed consent at the time of study enrollment. This study did not provide predictive genetic testing for genetic prion disease. If at-risk participants did not know their own genetic status, the clinical team was masked to their status. The research team performed *PRNP* genotyping on de-identified samples for research purposes only. For analyses conducted at MGH (assessments, post-LP survey, CSF total protein), participant data were collected and stored using REDCap^52^. To protect participant privacy, mutations carried by only one individual are grouped as “Other”, and the dates of participants’ second and third visits were scrambled by the addition of a normally distributed random variable (mean = 0, standard deviation = 2 weeks or 2 months, for second and third visits respectively). The study was originally designed to recruit 10 mutation carriers and 10 controls, a number expected to be sufficient to characterize test-retest reliability of CSF PrP as a descriptive statistic; enrollment was subsequently expanded as funding allowed. Of N=69 volunteers who contacted the study staff, N=26 were excluded due to LP contraindication (N=2), intercurrent illness (N=1), long travel distance for known mutation-negative participant (N=6), or non-responsiveness to follow-up (N=17). Of the 43 participants who enrolled and completed at least one visit, N=3 have been lost to follow-up after a first visit (N=1) or second visit (N=2), and N=4 are in touch but currently not participating due to intercurrent illness (N=2), LP contraindication (N=1), or unavailability to schedule another visit (N=1). An additional N=2 participants have missed one study visit due to pregnancy. Data points are omitted from analysis where missing due to missed visits or unsuccessful LPs.

### Assessments of cognitive, neuropsychiatric, motor and daily functioning

At each study visit, participants completed a battery of cognitive tests and standardized assessments of mood, neuropsychiatric symptoms, motor function, and activities of daily living. The cognitive battery consisted of standard paper and pencil neuropsychological measures including the Montreal Cognitive Assessment (MoCA)^18^, Verbal Fluency and Color Word Interference subtests within the Delis-Kaplan Executive Functioning System (D-KEFS)^53^, the Grooved Pegboard test^54^, Trailmaking Test Parts A and B^55^, and the DCTclock test, which is a digitized version of the standard clock drawing test^56^. Participants also completed computerized testing on an iPad consisting of the following subtests from the National Institute of Health (NIH) Toolbox Cognition Battery^57^: 1) Dimensional Change Card Sort, 2) Flanker Inhibitory Control and Attention Test, 3) Picture Sequence Memory Test, 4) List Sorting Working Memory Test, 5) Pattern Comparison Processing Speed Test, 6) Picture Vocabulary Test, 7) Reading Recognition Test and 8) Auditory Verbal Learning Test (Rey), with supplemental 20 minute delayed recall administered after completion of the toolbox. Raw scores obtained from cognitive measures were converted to standardized scores based on population-based normative data published for each test. Administered self-report questionnaires included the Beck Anxiety Inventory (BAI)^58^, Beck Depression Inventory (BDI)^59^, Measurement of Everyday Cognition-Short Version (ECog-12)^60,61^, Epworth Sleepiness Scale^62^, National Prion Monitoring Cohort MRC Scale^17^, Motor Aspects of Experiences of Daily Living section of the Movement Disorders Society-Unified Parkinson’s Disease Rating Scale (MDS-UPDRS M-EDL)^63^, and the clinician-administered Neuropsychiatric Inventory – Questionnaire (NPI-Q)^64^.

### Blood processing

Blood was collected in EDTA tubes, then centrifuged at 1000 rpm to separate plasma for aliquotting into 0.5 mL aliquots. DNA was extracted from whole blood collected on the first study visit. All samples were codified for analysis. Genotypes were used for research purposes only. *PRNP* single nucleotide variants were identified at the Broad Institute Genomics Platform using a custom targeted capture platform developed by Twist Bioscience combined with deep Illumina sequencing. These genotypes were then confirmed, and the presence or absence of octapeptide repeat insertions determined, using a previously described sequencing and gel analysis protocol^65^ implemented by Genewiz and/or Quintarabio. Briefly, this analysis uses primers Int5: 5’-TgCATgTTTTCACgATAgTAACgg-3’, DG2: 5’-gCAgTCATTATggCgAACCTTggCTg-3’, and 3’Sal: 5’-gTACTgAggATCCTCCTCATCCCACTATCAggAAgA-3’. A DG2/3’Sal PCR product (wild-type: 804bp) is subjected to Sanger sequencing while a DG2/Int5 PCR product (wild-type: 464bp) is run on a 2% agarose gel to identify large indels.

### Lumbar puncture and CSF processing

The lumbar puncture (LP) for CSF collection was performed using a standardized protocol with a 24G atraumatic Sprotte needle. The time of day for LP was kept consistent across subjects and 20 mL CSF was collected per subject where possible. Following collection, CSF was handled uniformly according to an established protocol designed to minimize PrP loss to plastic through measures including i) highly controlled and minimized plastic exposure, ii) uniform storage in aliquots no smaller than 40 μL, and iii) addition of 0.03% CHAPS detergent to a subset of CSF to maintain PrP solubility^16^. Samples were then frozen and banked at the Broad Institute where they were stored at −80°C until analysis. CSF aliquots containing 0.03% CHAPS were used for PrP quantification by ELISA; neat CSF aliquots with no additive were used for T-tau ELISA, NfL ELISA, and RT-QuIC. Because some LPs were anomalous or unsuccessful, for some participants CHAPS CSF, neat CSF, or both were not available. These individuals were excluded from the corresponding analyses. All analyses were performed by researchers blinded to participant identity and carrier/control status.

### Samples from symptomatic prion disease patients

N=26 anonymized pre-mortem cerebrospinal fluid samples from symptomatic sporadic (N=22) and genetic (N=4, all E200K) prion disease cases collected between 2001 and 2017 were shared by the Australian National Creutzfeldt-Jakob Disease Registry (ANCJDR). All cases were autopsy-confirmed as prion disease, except for N=2 genetic cases, which due to the presence of the mutation are highly likely to have been prion disease. Originally received in ∼0.5 mL aliquots, samples were thawed once following receipt, aliquotted to 100 μL volume, refrozen at −80°C, and thereafter thawed only immediately preceding experimental use. Due to sample volume limitations, not all positive controls were utilized in all assays.

### Post-LP survey

Following each LP, participants completed a brief survey that we designed to assess the experience, either on paper or via iPad. They were asked whether they had previously had an LP, and if so, how many. Participants were then asked to mark an X on a 14-cm Likert-type scale to indicate 1) their level of anxiety before the LP procedure, and 2) their current feelings at the prospect of a future LP. In both cases, the response was marked on a continuous spectrum bounded by the two extremes of “Not anxious at all” and “Extremely anxious.” Responses were normalized to the full length of the scale.

### PrP ELISA

PrP levels were quantified at the Broad Institute using the BetaPrion Human ELISA assay, according to the manufacturer’s instructions (AnalytikJena, Leipzig, Germany). As described previously,^16^ to maintain PrP in solution, CSF samples used for this analysis were handled with close attention to uniformity, and were spiked with 0.03% CHAPS detergent immediately after collection. All samples were diluted 1:50 in blocking buffer (0.05% Tween, 5% BSA, 1x PBS) and assayed in duplicate, with samples from the same individual co-located on the same plate to facilitate comparison. Following termination of the colorimetric development reaction, absorbance per well was measured at 450 nm as well as at 620 nm for background subtraction using a FLUOStar Optima absorbance plate reader, then fit to an internal standard curve to generate PrP concentrations in ng/mL. The operator was blinded to mutation status.

### Total tau ELISA (Broad Institute and University of Gothenburg)

CSF T-tau was measured using the INNOTEST hTau Ag ELISA kit (Fujirebio, Malvern, PA, USA and Ghent, Belgium) according to the manufacturer’s instructions. Study samples were diluted 1:4; positive control symptomatic prion disease samples were diluted 1:10. All samples were assayed in duplicate with samples from the same individual co-located on the same plate to facilitate comparison. Following termination of the colorimetric development reaction, absorbance per well was measured at 450 nm as well as at 620 nm for background subtraction using a FLUOStar Optima absorbance plate reader, then fit to an internal standard curve. The operator was blinded to mutation status.

### NfL ELISA (Broad Institute)

CSF NfL was measured using the NF-light RUO ELISA (Uman Diagnostics, IBL International, Umea, Sweden) according the manufacturer’s instructions. Study samples were diluted 1:2; positive control symptomatic prion disease samples were diluted 1:5. All samples were assayed in duplicate with samples from the same individual co-located on the same plate to facilitate comparison. Following termination of the colorimetric development reaction, absorbance per well was measured at 450 nm as well as at 620 nm for background subtraction using a FLUOStar Optima absorbance plate reader, then fit to an internal standard curve. The operator was blinded to mutation status.

### NfL ELISA (University of Gothenburg)

Following CSF collection and processing as described above, uniformly handled 0.5 mL CSF aliquots with no additive were stored at - 80°C until shipment on dry ice to the University of Gothenburg for analysis. CSF neurofilament light (NfL) was measured using an in-house developed ELISA as previously described^66^.

### Simoa analysis of plasma

Following blood processing as described above, 0.5 mL plasma aliquots were stored at - 80°C until shipment on dry ice to the University of Gothenburg for analysis. Plasma NfL and total tau levels were measured using the Single molecule array (Simoa) HD-1 Analyzer (Quanterix, Billerica, MA, USA). For T-tau, the commercially available Tau 2.0 kit was used according to the manufacturer’s instructions (Quanterix). For NfL, a previously described in house Simoa assay was used^24^. Calibrators were run in duplicate and obvious outlier calibrator replicates were masked before curve fitting. Samples were run in singlicate with 4-fold dilution. Two quality control samples were run in duplicate at the beginning and end of each run.

### RT-QuIC (Syrian hamster prion protein substrate)

The real-time quaking induced conversion (RT-QuIC) assay was performed according to an established RT-QuIC protocol for detection of prion seeds in CSF^25^ that is widely used for diagnosis of symptomatic prion disease patients. Briefly, truncated recombinant Syrian hamster prion protein (SHaPrP 90-230) was purified from *E. coli* according to established protocols^67^, then frozen at −80°C following determination of concentration by NanoDrop. On the day of use, PrP was thawed and centrifuged at 5,000 × g for 5 minutes at 4C in a PALL 100 kDa filter tube. 80 μL of reaction mix and 20 μL of CSF were combined in each well of a black 96-well plate with a clear bottom (Nunc) with final concentrations as follows: 300 mM NaCl, 10 mM phosphate buffer, 1 mM EDTA, 10 uM thioflavin T, 0.002% SDS, 1 mg/mL SHaPrP. All samples were loaded in quadruplicate with each plate containing negative control CSF (healthy mutation-negative individuals) and positive control CSF (symptomatic prion disease patients). After sealing (Nalgene Nunc International sealer), plates were incubated in a BMG FLUOstar Optima plate reader at 55°C for 40h with continuous cycles of 60 s shaking (700 rpm, double-orbital) and 60 s rest, and ThT fluorescence measurements every 45 min (excitation 450 nm, emission 480 nm, bottom read.) Following termination of the experiment, fluorescence readings were merged per well to generate kinetic curves, and the threshold for a positive well was set as the mean value of all negative wells plus 10 standard deviations. A sample was considered overall positive if at least two of four replicates crossed this threshold. The operator was blinded to mutation status.

### RT-QuIC (bank vole prion protein substrate)

For the BvPrP alternative protocol, RT-QuIC was performed according to the protocol described by Orru et al^28^, with the key modification that 20 μL CSF seed was used per well, rather than 2 μL brain homogenate seed. The total reaction volume remained 100 μL per well. As above, full-length recombinant bank vole prion protein (BvPrP 23-230) was purified from *E. coli* according to established protocols, then frozen at - 80°C following determination of concentration by NanoDrop. On the day of use, PrP was thawed and centrifuged at 5,000 × *g* for 5 minutes at 4°C in a PALL 100 kDa filter tube. 80 μL of reaction mix and 20 μL of CSF were combined in each well of a black 96-well plate with a clear bottom (Nunc) with final concentrations as follows: 300 mM NaCl, 10 mM phosphate buffer, 1 mM EDTA, 10 μM thioflavin T, 0.002% SDS, 1 mg/mL BvPrP. All samples were loaded in quadruplicate with each plate containing negative control CSF (healthy mutation-negative individuals) and positive control CSF (symptomatic prion disease patients). After sealing (Nalgene Nunc International sealer), plates were incubated in a BMG FLUOstar Optima plate reader at 42°C for 50h with continuous cycles of 60 s shaking (700 rpm, double-orbital) and 60 s rest, and ThT fluorescence measurements every 45 min (excitation 450 nm, emission 480 nm, bottom read.) Following termination of the experiment, fluorescence readings were merged per well to generate kinetic curves, and the threshold for a positive well was set as the mean value of all negative wells plus 10 standard deviations. A sample was considered overall positive if at least two of four replicates crossed this threshold. The operator was blinded to mutation status.

## Data Availability

R source code for this study is available in a public repository: https://github.com/ericminikel/mgh_prnp_fluid_biomarkers De-identified data will be made available to qualified investigators upon request.

## ACKNOWLEDGMENTS

This study was supported by Prion Alliance, CJD Foundation (the Michael H. Cole, Cheryl Molloy, José A. Piriz and Sonia E. Piriz, Jeffrey A. Smith, and Mercies in Disguise Memorial Grants), the Broad Institute (Broad Next Gen Fund and direct donations to Prions@Broad), the National Science Foundation (GRFP 2015214731 to SMV), the National Institutes of Health (F31 AI122592 to EVM and R21 TR003040 to SEA), and an anonymous organization. We thank Brendan Blumenstiel and Anna Koutoulas for assistance with DNA sequencing. We are grateful to all of the volunteers who participated in this research study.

## AUTHOR CONTRIBUTIONS

Conceived and designed the study: SMV, EVM, SEA

Performed the experiments: SMV, EVM, VJW, BCC, AJM, CDW, AB, BAT, CKN, HD, DU, JG

Provided key samples and reagents: CS, SJC, IL

Supervised the research: CS, SJC, KB, HZ, SEA

Drafted the manuscript: SMV, EVM

Reviewed, edited, and approved the final manuscript: All authors

## SUPPLEMENT

**Figure S1:**
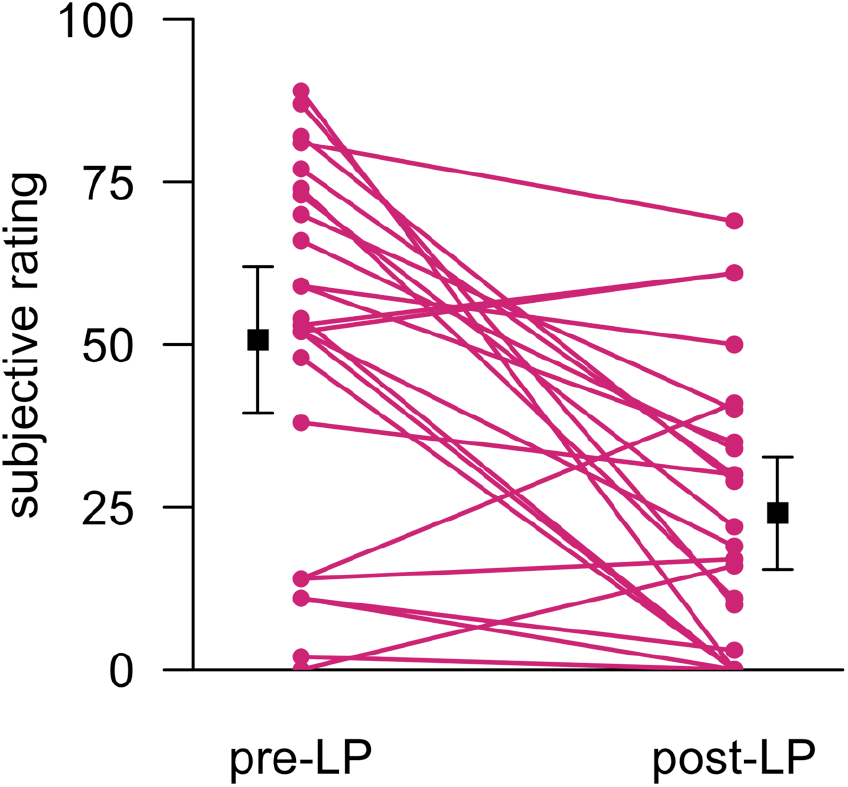
Pre- and post-procedure anxiety in participants experiencing their first lumbar puncture. Following the lumbar puncture procedure, participants were asked to quantitatively rank both their anxiety before the procedure and their post-procedure anxiety when contemplating a future LP, using a Likert-type scale. Responses are normalized to the length of the scale and shown only for first study visits, for N=24 individuals who had not undergone a previous LP.

**Figure S2:**
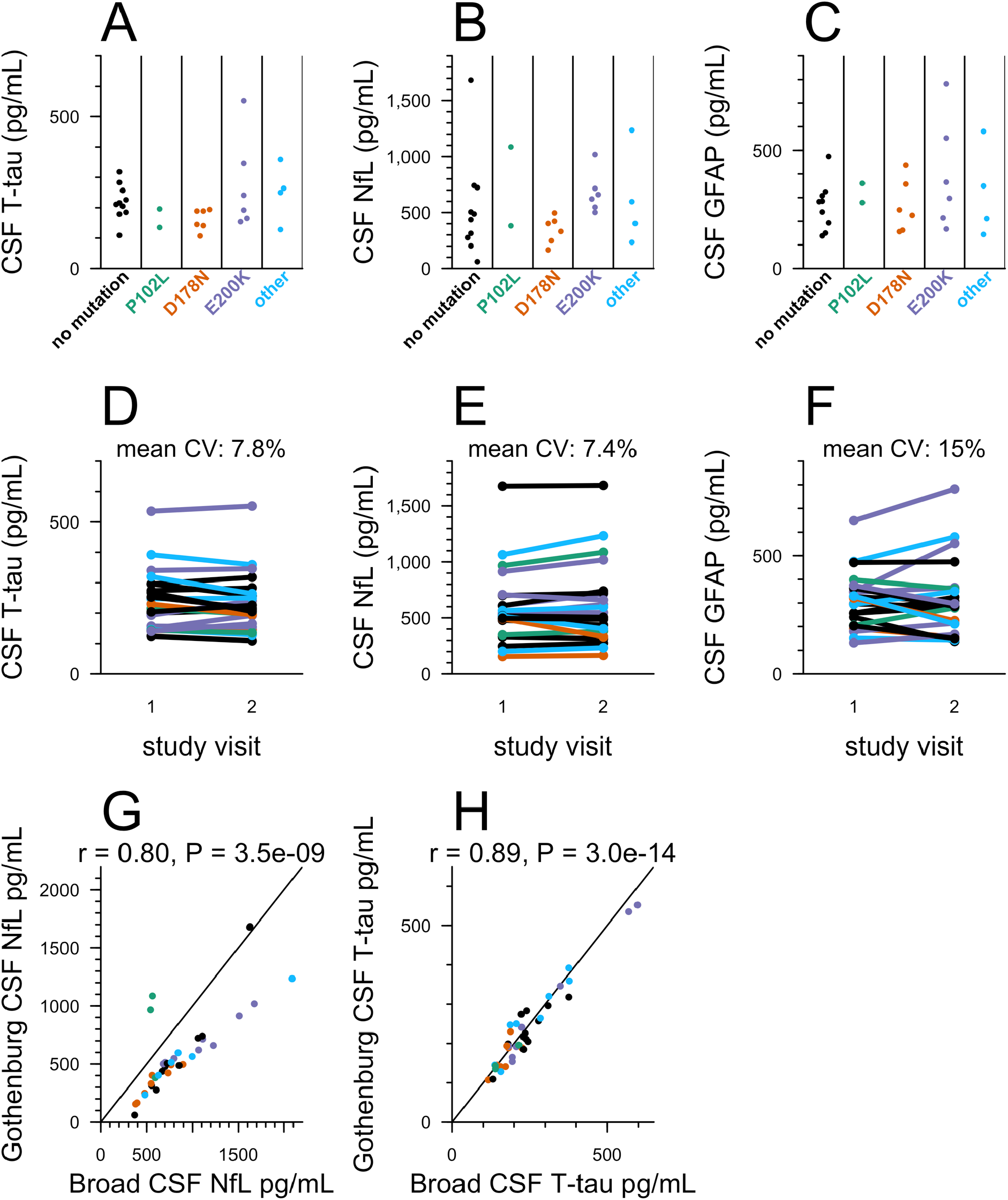
Markers of neuronal damage in carrier and control CSF (University of Gothenburg). CSF **A)** T-tau and **B)** NfL and C) glial fibrillary acidic protein (GFAP) were measured by ELISA for N=28 participants who had made at least one study visit, for whom genotypes were available at time of analysis, and where appropriate CSF aliquots were available. For each participant included, samples were taken from the most recent visit at time of analysis. The operator was masked to mutation status. Dots represent means, and line segments 95% confidence intervals, of measurements within dynamic range with 2 technical replicates each. **D-F)** For N=22 participants who had completed two visits at time of analysis, both total tau and NfL were measured by ELISA for both visits to assess within-subject variability over a 2-4 month timeframe. E-F) Correlation between results obtained from two independent ELISA analyses at different sites (performed using different assays for NfL, and the same commercial assay for T-tau; see Methods) for N=36 samples from N=27 individuals (G) and N=39 samples from N=27 individuals (H).

**Figure S3:**
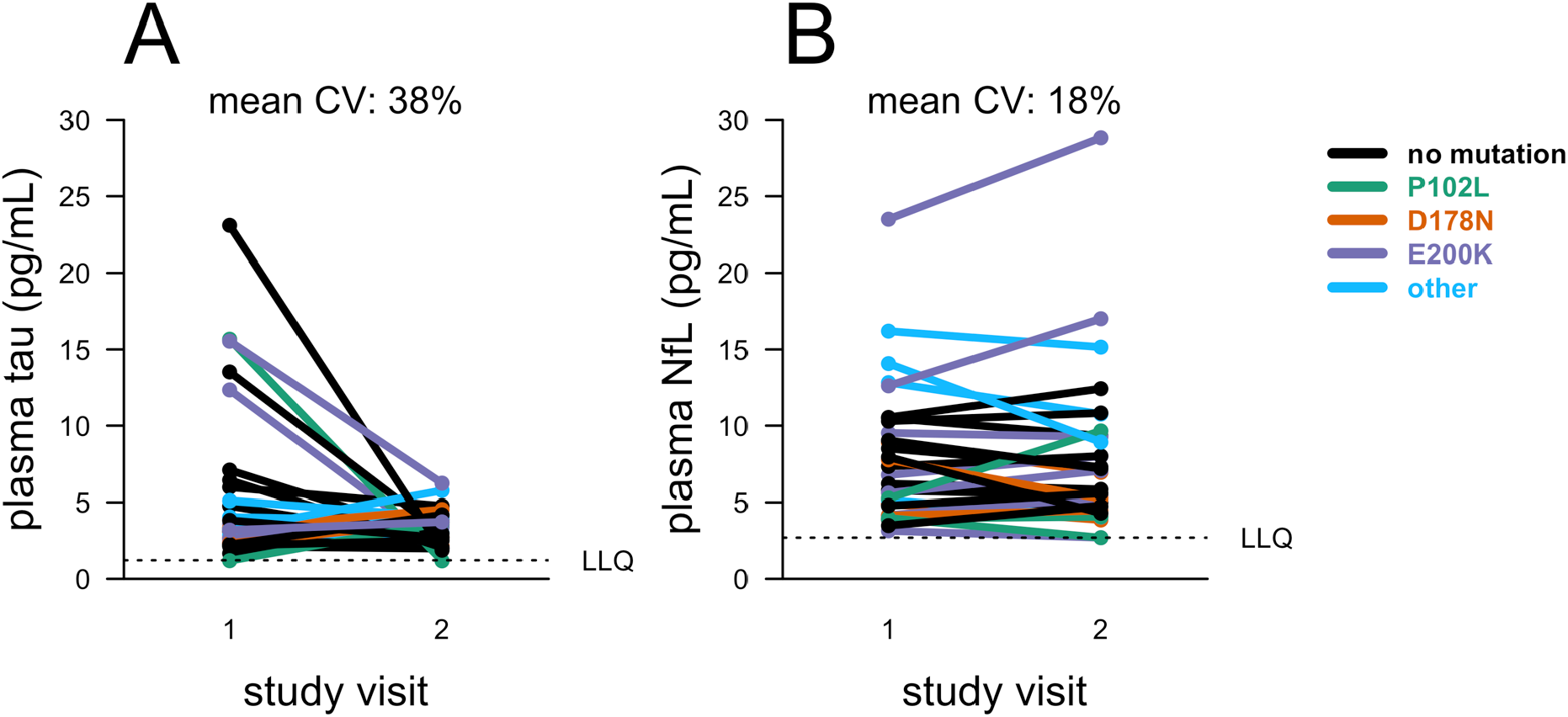
Short-term test-retest stability of markers of neuronal damage in carrier and control plasma (University of Gothenburg). Plasma T-tau and NfL levels were measured by Quanterix Simoa assay. For N=30 participants (T-tau) or N=31 participants (NfL) who have made at least two study visits, samples were taken from the initial two study visits, completed 2-4 months apart. Dots represent singlicate measurements. For participants with exactly two visits, the NfL measurement from the second visit displayed here is also displayed in Figure 3.

**Figure S4:**
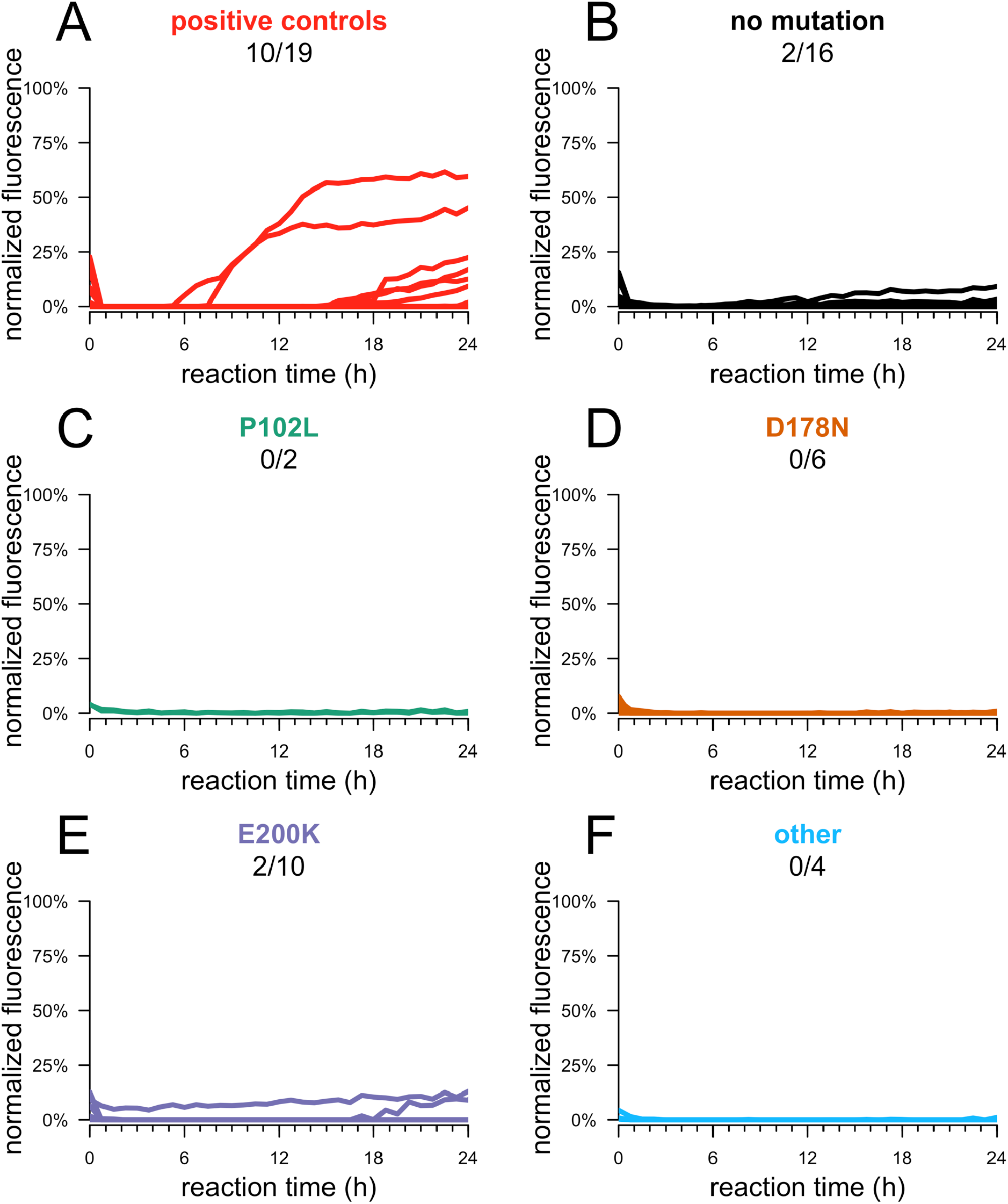
RT-QuIC results with recombinant bank vole PrP. RT-QuIC was performed on CSF from 39 participants who have made at least one study visit, selected to match the samples used in the SHaPrP RT-QuIC analysis (Figure 4). RT-QuIC was performed following a published protocol for RT-QuIC using BvPrP substrate, intended for prion detection in brain homogenate. Reagent concentrations were adjusted to accommodate 20 μL CSF in a final reaction volume of 100 μL. N=22 prion disease cases or N=39 MGH study participants were assayed, with each reaction run in quadruplicate. Kinetic curves – normalized thioflavin T (ThT) fluorescence (y axis) vs. time in hours (x axis) – are shown for each replicate.

**Table S1.**
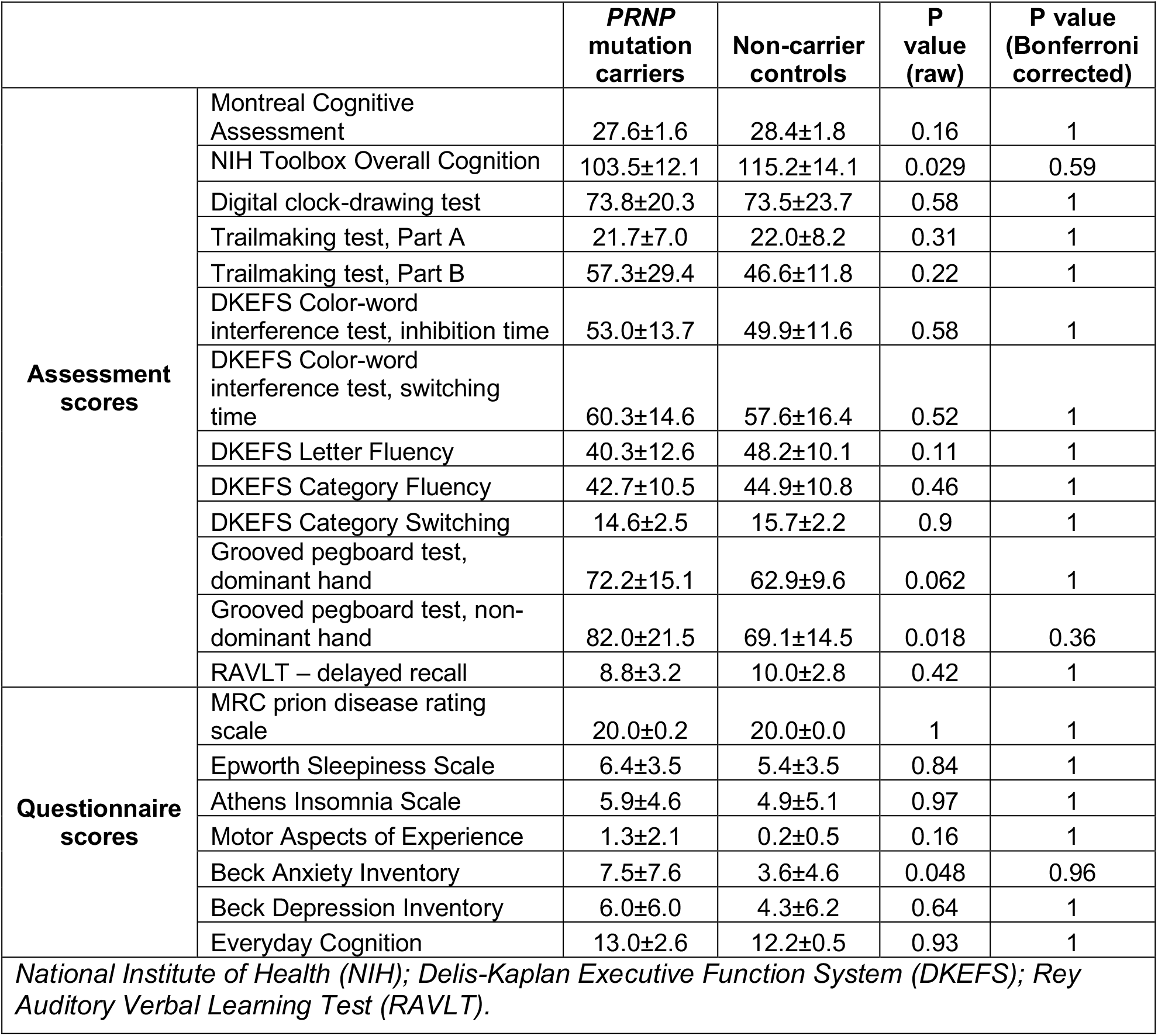
Measures of cognitive, psychiatric, motor and daily functioning in all MGH study participants. Scores are averaged across group (carrier vs. non-carrier) at first completed study visit. P values are from two-sided Kolmogorov-Smirnov tests. Bonferroni corrected p values account for a multiple testing burden of N=20 for all measures shown. Raw scores are provided according to the methods standard for each test, with the exception of the NIH Toolbox overall cognition composite score, and the digital clock drawing test, where digitally recorded features were compiled into a composite score as previously described^56^.

**Table S2.** Additional measures of cognitive, psychiatric, motor and daily functioning for one RT-QuIC positive MGH study participant. Visits were separated by two months. Normative scores (based on normative data matched by age, gender, and education) are provided according to the published standard for each test.

|  |  | Visit 1 |  | Visit 2 |  | Reliable Change Interval |
| --- | --- | --- | --- | --- | --- | --- |
| Assessment scores |  | Raw | Standard (%ile) | Raw | Standard (%ile) |  |
| | Montreal Cognitive Assessment | 27 | -- | 25 | -- | $\pm 2.47$ |
| | NIH Toolbox Overall Cognition | 109 | -- | 104 | -- | $\pm 10.17$ |
| | Digital clock drawing test | 51 | -- | 56 | -- | $\pm 16.04$ |
| | Trailmaking test, Part A | 40 | 0.11 (54) | 51 | -0.67 (25) | $\pm 8.49^*$ (↓) |
| | Trailmaking test, Part B | 178 | -1.75 (4) | 120 | -0.45 (33) | $\pm 13.17^*$ (↑) |
| | DKEFS Color-word interference test, inhibition time | 63 | 12 (75) | 70 | 10 (50) | $\pm 12.80$ |
| | DKEFS Color-word interference test, switching time | 86 | 9 (37) | 73 | 11 (63) | $\pm 11.90^*$ (↑) |
| | DKEFS Letter Fluency | 43 | 12 (75) | 37 | 11 (63) | $\pm 16.11$ |
| | DKEFS Category Fluency | 38 | 12 (75) | 36 | 11 (63) | $\pm 14.27$ |
| | DKEFS Category Switching | 12 | 10 (50) | 14 | 11 (63) | $\pm 3.37$ |
| | Grooved pegboard test, dominant hand | 122 | -2.14 (2) | 131 | -2.60 (<1) | $\pm 11.42$ |
| | Grooved pegboard test, non-dominant hand | 157 | -2.67 (<1) | 155 | -2.59 (<1) | $\pm 13.59$ |
| | RAVLT– delayed recall | 6 | -0.42 (34) | 6 | -0.42 (34) | $\pm 3.20$ |
| Questionnaire scores | Epworth Sleepiness Scale | 3 | -- | 2 | -- | -- |
|  | Athens Insomnia Scale | 7 | -- | 1 | -- | -- |
|  | Motor Aspects of Experience | 1 | -- | 0 | -- | -- |
|  | Beck Anxiety Inventory | 9 | -- | 5 | -- | -- |
|  | Beck Depression Inventory | 4 | -- | 6 | -- | -- |
|  | Everyday Cognition | 12 | -- | 12 | -- | -- |

*Delis-Kaplan Executive Function System (DKEFS); Rey Auditory Verbal Learning Test (RAVLT); Standard scores for DKEFS subtests are presented as scaled scores (M=10, SD=3); Standard scores for Trailmaking test, Grooved pegboard, and RAVLT are presented as z-scores (M=0, SD=1). The reliable change interval was calculated based on the standard deviation of the residuals predicting time 2 scores from time 1 scores (using a two-tailed 90% confidence interval) within the non-carrier control group. After raw scores were standardized based on published age and gender norms for each test, this individual performed within the average range for the majority of cognitive measures administered at both time 1 and time 2. The notable exception is the grooved pegboard test where this individual's performance consistently fell within the impaired range for both dominant and non-dominant hands, and likely represents an area of relative weakness that is consistent over time. Although performance on Trailmaking Test Part B was also impaired at time point 1, performance significantly improved across the retest interval and the score was within the average range at time 2. \*Tests falling outside of the confidence interval of reliable change between visits with accompanying arrows indicating whether performance at visit 2 was better (↑) or worse (↓) than performance at visit 1.*

## STROBE Statement

### checklist of items that should be included in reports of observational studies

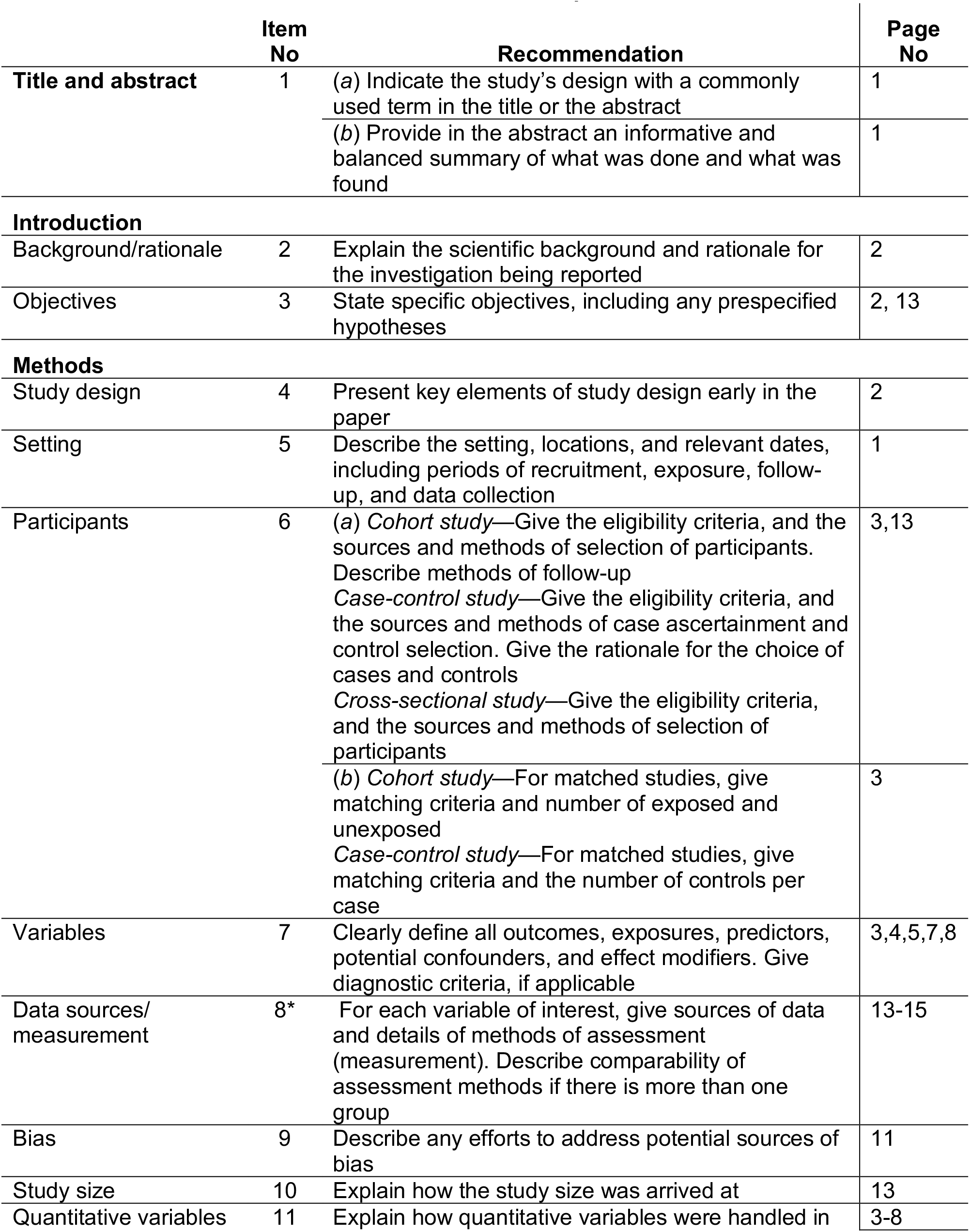
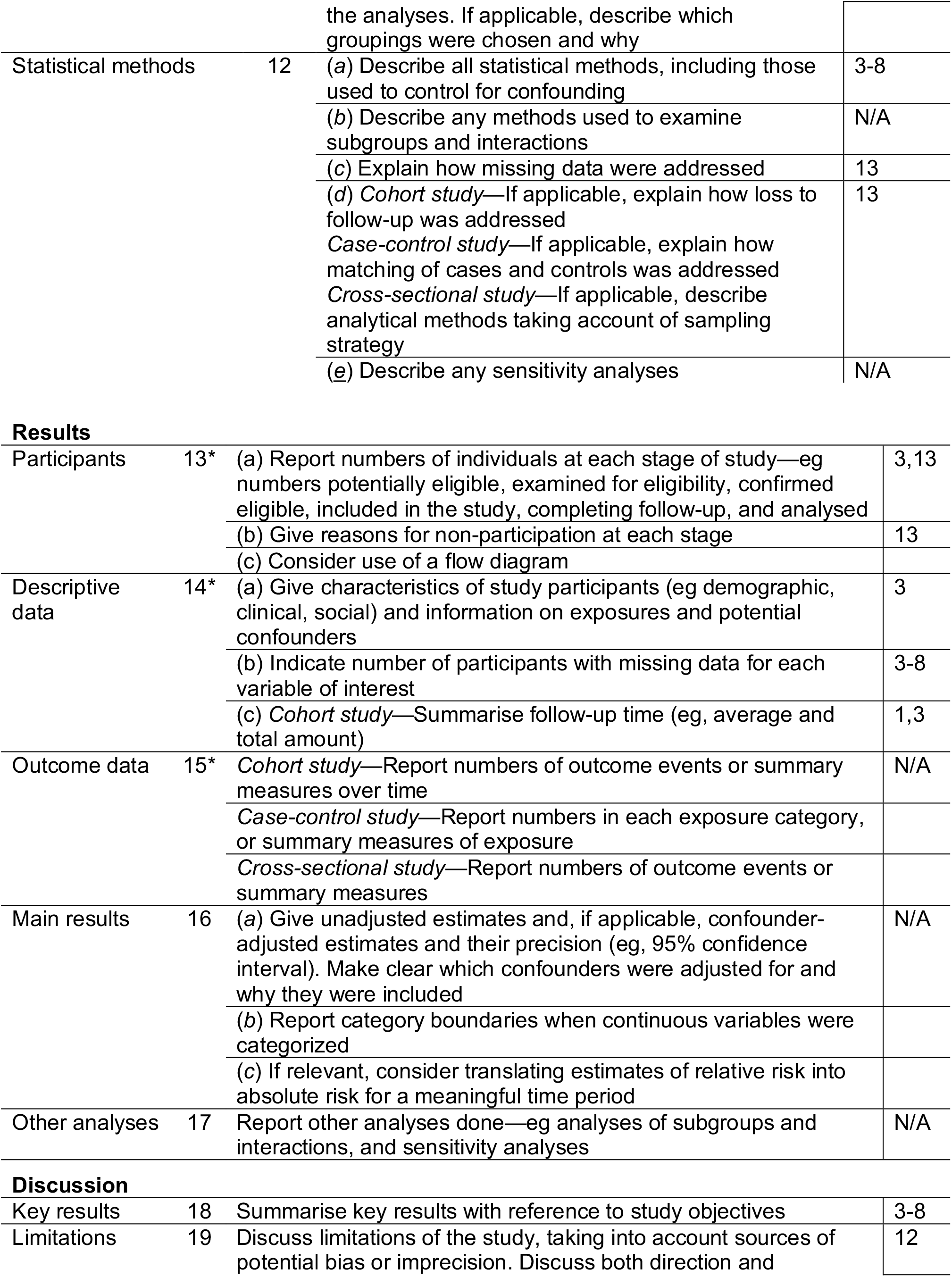
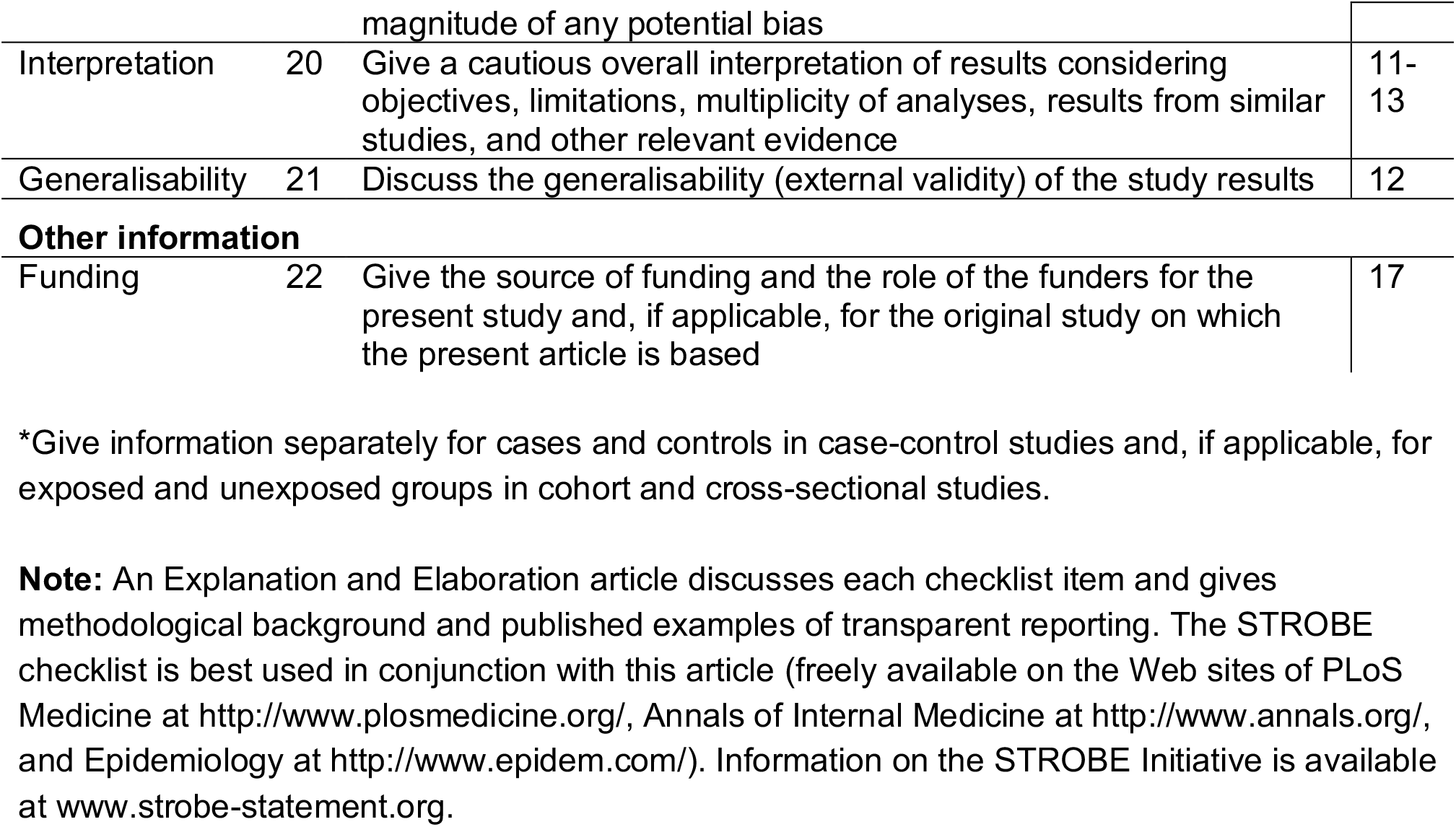

|  | Item No | Recommendation | Page No |
| --- | --- | --- | --- |
| Title and abstract | 1 | (a) Indicate the study’s design with a commonly used term in the title or the abstract | 1 |
|  |  | (b) Provide in the abstract an informative and balanced summary of what was done and what was found | 1 |
| Introduction |  |  |  |
| Background/rationale | 2 | Explain the scientific background and rationale for the investigation being reported | 2 |
| Objectives | 3 | State specific objectives, including any prespecified hypotheses | 2, 13 |
| Methods |  |  |  |
| Study design | 4 | Present key elements of study design early in the paper | 2 |
| Setting | 5 | Describe the setting, locations, and relevant dates, including periods of recruitment, exposure, follow-up, and data collection | 1 |
| Participants | 6 | (a) Cohort study—Give the eligibility criteria, and the sources and methods of selection of participants. Describe methods of follow-up<br>Case-control study—Give the eligibility criteria, and the sources and methods of case ascertainment and control selection. Give the rationale for the choice of cases and controls<br>Cross-sectional study—Give the eligibility criteria, and the sources and methods of selection of participants | 3,13 |
|  |  | (b) Cohort study—For matched studies, give matching criteria and number of exposed and unexposed<br>Case-control study—For matched studies, give matching criteria and the number of controls per case | 3 |
| Variables | 7 | Clearly define all outcomes, exposures, predictors, potential confounders, and effect modifiers. Give diagnostic criteria, if applicable | 3,4,5,7,8 |
| Data sources/measurement | 8* | For each variable of interest, give sources of data and details of methods of assessment (measurement). Describe comparability of assessment methods if there is more than one group | 13-15 |
| Bias | 9 | Describe any efforts to address potential sources of bias | 11 |
| Study size | 10 | Explain how the study size was arrived at | 13 |
| Quantitative variables | 11 | Explain how quantitative variables were handled in the analyses. If applicable, describe which groupings were chosen and why | 3-8 |
| Statistical methods | 12 | (a) Describe all statistical methods, including those used to control for confounding | 3-8 |
|  |  | (b) Describe any methods used to examine subgroups and interactions | N/A |
|  |  | (c) Explain how missing data were addressed | 13 |
|  |  | (d) *Cohort study*—If applicable, explain how loss to follow-up was addressed<br>*Case-control study*—If applicable, explain how matching of cases and controls was addressed<br>*Cross-sectional study*—If applicable, describe analytical methods taking account of sampling strategy | 13 |
|  |  | (e) Describe any sensitivity analyses | N/A |
| Participants | 13* | (a) Report numbers of individuals at each stage of study—eg numbers potentially eligible, examined for eligibility, confirmed eligible, included in the study, completing follow-up, and analysed | 3,13 |
|  |  | (b) Give reasons for non-participation at each stage | 13 |
|  |  | (c) Consider use of a flow diagram |  |
| Descriptive data | 14* | (a) Give characteristics of study participants (eg demographic, clinical, social) and information on exposures and potential confounders | 3 |
|  |  | (b) Indicate number of participants with missing data for each variable of interest | 3-8 |
|  |  | (c) *Cohort study*—Summarise follow-up time (eg, average and total amount) | 1,3 |
| Outcome data | 15* | *Cohort study*—Report numbers of outcome events or summary measures over time | N/A |
|  |  | *Case-control study*—Report numbers in each exposure category, or summary measures of exposure |  |
|  |  | *Cross-sectional study*—Report numbers of outcome events or summary measures |  |
| Main results | 16 | (a) Give unadjusted estimates and, if applicable, confounder-adjusted estimates and their precision (eg, 95% confidence interval). Make clear which confounders were adjusted for and why they were included | N/A |
|  |  | (b) Report category boundaries when continuous variables were categorized |  |
|  |  | (c) If relevant, consider translating estimates of relative risk into absolute risk for a meaningful time period |  |
| Other analyses | 17 | Report other analyses done—eg analyses of subgroups and interactions, and sensitivity analyses | N/A |
| Key results | 18 | Summarise key results with reference to study objectives | 3-8 |
| Limitations | 19 | Discuss limitations of the study, taking into account sources of potential bias or imprecision. Discuss both direction and magnitude of any potential bias | 12 |
| Interpretation | 20 | Give a cautious overall interpretation of results considering objectives, limitations, multiplicity of analyses, results from similar studies, and other relevant evidence | 11-13 |
| Generalisability | 21 | Discuss the generalisability (external validity) of the study results | 12 |
| **Other information** |  |  |  |
| Funding | 22 | Give the source of funding and the role of the funders for the present study and, if applicable, for the original study on which the present article is based | 17 |

\*Give information separately for cases and controls in case-control studies and, if applicable, for exposed and unexposed groups in cohort and cross-sectional studies.
**Note:** An Explanation and Elaboration article discusses each checklist item and gives methodological background and published examples of transparent reporting. The STROBE checklist is best used in conjunction with this article (freely available on the Web sites of PLoS Medicine at http://www.plosmedicine.org/, Annals of Internal Medicine at http://www.annals.org/, and Epidemiology at http://www.epidem.com/). Information on the STROBE Initiative is available at www.strobe-statement.org.

